# Seed-based connectivity of ventral tegmental area correlates with decreased physical fitness in schizophrenia

**DOI:** 10.1101/2022.06.27.22276810

**Authors:** Lara Hamzehpour, Tamara Bohn, Lucia Jaspers, Oliver Grimm

## Abstract

**Background:** Overweight and decreased physical fitness are highly prevalent in schizophrenia, represent a major risk factor for cardio-vascular diseases and decrease the patients’ life expectancies. It is thus important to understand the underlying mechanisms that link psychopathology and weight gain. We hypothesize that the dopaminergic reward system plays an important role in this.

**Methods:** We analyzed the seed-based functional connectivity (FC) of the ventral tegmental area (VTA) in a group of schizophrenic patients (n = 32) and age- as well as gender matched healthy controls (n = 27). We then correlated the resting-state results with physical fitness parameters, obtained in a fitness test, and psychopathology.

**Results:** The seed-based connectivity analysis revealed decreased functional connections between the VTA and the anterior cingulate cortex (ACC), as well as the dorsolateral prefrontal cortex and increased functional connectivity between the VTA and the middle temporal gyrus in patients compared to healthy controls. The decreased FC between the VTA and the ACC of the patient group could further be associated with increased body fat and negatively correlated with the overall physical fitness. We found no significant correlations with psychopathology.

**Conclusion:** Although we did not find significant correlations with psychopathology, we could link decreased physical fitness and high body fat with dysconnectivity between the VTA and the ACC in schizophrenia. These findings demonstrate that a dysregulated reward system is not just responsible for symptomatology in schizophrenia but is also involved in comorbidities and could pave the way for future lifestyle therapy interventions.

## 1 Introduction

As one of the most debilitating psychiatric disorders, schizophrenia affects about 1 percent of the world’s population. The onset usually occurs in late adolescence and symptoms typically remain throughout life [1]. Schizophrenia is characterized by hallucinations and delusions, referred to as positive symptoms as well as severe cognitive and affective impairments, known as negative symptoms [2]. While this places a heavy burden on patients, leading to social isolation and the inability to cope with everyday life [3], schizophrenia - like other mental illnesses - is further accompanied by weight gain and decreased physical fitness [4,5]. Strassnig and colleagues [5], for example, examined the cardiorespiratory fitness of a male schizophrenic cohort, revealing that 98.3% of the 117 participants scored below population standards. Complicated by obesity, low cardio-respiratory fitness represents a major risk factor for cardio-metabolic disturbances, referred to as metabolic syndrome [6-9]. It usually comprises symptoms like central and abdominal adiposity, insulin resistance, dyslipidaemias, glucose intolerance and hypertension. Metabolic syndrome, in turn, is predictive of severe physical comorbidities, such as type II diabetes or cardiovascular disease. With regard to schizophrenia, obesity is three times as prevalent in patients compared to the general population [10], with obese patients having a higher risk to developing cardio-vascular diseases compared to obese but otherwise healthy individuals [11]. Numerous studies could show that, besides suicide, cardio-metabolic diseases represent one of the major causes of premature death in patients suffering from schizophrenia with their life expectancies being reduced by 10 to 25 years compared to the general population [12].

The reasons for the increased risk of patients to be in poorer physical conditions are diverse and complex. First, there are genetic risk factors [13] and the cardio-vascular side-effects of antipsychotic medication [10,13]. Second, patients have the tendency to maintain a rather unhealthy lifestyle, characterized by increased smoking rates, poor diets and physical inactivity. Thirdly, the predisposition of patients with schizophrenia to gain weight is related to the disorder’s neurobiology itself [8,14].

Schizophrenia’s neurobiology is characterized by a dysregulation of dopaminergic neurotransmission in the mesocorticolimbic circuit of the brain [15-18]. These dopaminergic neurons primarily originate from the midbrain, especially the ventral tegmental area (VTA), and project to regions like the prefrontal cortex, ventral pallidum and nucleus accumbens, which regulate reward and motivational processes [18-21]. Consequently, disorder-related dopaminergic impairments are responsible for a variety of symptoms, including cognitive deficits, psychosis and altered reward processing [17,22]. The reward circuit has further become the subject of schizophrenia research in many neuroimaging studies. This way, striatal hypoactivation during reward anticipation could be identified as a phenotype of schizophrenia [22,23] and resting state data revealed functional dysconnectivity of the reward system to correlate with symptom severity in patients [24,25].

There is further evidence that VTA dopamine projections are implicated in the coordination of energy balance, which comprises the regulation of food intake and voluntary physical activity [26-28]. As disturbances in any of these processes would lead to an increased intake of palatable foods and less motivation to become physically active [29], we hypothesize that illness-related alterations in the dopaminergic reward system might be responsible for both symptom severity and decreased physical fitness of schizophrenic patients.

To our knowledge no study to date has attempted to establish a link between alterations in the reward system and physical fitness in schizophrenia. Therefore, we aim to demonstrate that functional dysconnectivity in schizophrenic patients correlates with increased psychopathology and reduced physical fitness compared to healthy individuals, using the VTA - the source of corticostriatal dopamine neurons - as our seed region in the seed-to-voxel resting-state analysis.

## 2 Materials and Methods

### 2.1 Participants

30 patients and 30 healthy participants were matched as to gender and age, with a mean age of 35.1 for the patient group and 34.8 for the healthy subject group. Patients were mainly recruited from the department of Psychiatry, Psychosomatic Medicine and Psychotherapy of the university clinic in Frankfurt am Main, Germany and psychiatric institutions of surrounding areas via flyers and web pages. Psychiatric diagnoses were based on DSM-V criteria and established by registered psychiatrists responsible for the patients’ treatment. Patients were included in the study if the following criteria were met: 1) diagnosis of schizophrenia ; 2) no abuse of cocaine or amphetamines within the last 2 weeks; 3) no intake of benzodiazepines within the last two weeks; 4) history of stable medication for at least 4 weeks; 5) no physical impairments that would hinder the performance in the fitness test; 6) no history of neurological disorders; 7) no current alcohol or drug abuse; 7) no magnetic resonance imaging (MRI) contraindications. Inclusion criteria for healthy participants were: 1) no history of psychiatric disorders; 2) no physical impairments that would hinder the performance in the fitness test; 3) no history of neurological disorders; 4) no current alcohol or drug abuse; 5) no MRI contraindications.

### 2.2 Psychopathology Assessment

To assess psychopathology and especially negative symptoms in both groups, we utilized the Chapman Scale for Physical and Social Anhedonia (PAS, SAS) [30], the World Health Organization Disability Assessment Schedule (WHODAS) [31] and the Calgary Depression Scale for Schizophrenia (CDSS) [32]. Clinical symptoms within the patient group were additionally recorded by means of the standardized Positive and Negative Syndrome Scale (PANSS) interview [33]. All interviews were conducted by trained raters.

Demographics and clinical characteristics of the study sample are summarized in Table 1. To decide whether group comparisons could be calculated using parametric (two-sample t-test) or non-parametric tests (Wilcoxon-rank-sum-test) [34], we first examined if the different variables were normally distributed using the Shapiro-Wilk-Test [35].

**Table 1.** Sample characteristics with two-sample t-test and Wilcoxon test results, displaying group differences between patients and healthy controls.

| characteristics | SZ | mean | SD | HC | mean | SD |
| --- | --- | --- | --- | --- | --- | --- |
| males | 22 |  |  | 18 |  |  |
| females | 10 |  |  | 9 |  |  |
| age [years] | 32 | 35.1 | $\pm 12.3$ | 27 | 34.8 | $\pm 12.2$ |
| CPZeq | | 602.2 | $\pm 465.4$ | | | |
| PANSS | 29 | 30.6 | $\pm 6.4$ | | | |
| PAS | | 13.9* | $\pm 8.5$ | 27 | 8.7* | $\pm 5.9$ |
| SAS | | 13.6* | $\pm 8.5$ | | 9* | $\pm 5.9$ |
| WHODAS | | 26.9** | $\pm 15.5$ | | 9.2** | $\pm 9.7$ |
| CDSS | | 3.7* | $\pm 2.8$ | | 2* | $\pm 3.2$ |
| BMI | 29 | 26.9* | $\pm 4.1$ | 27 | 25* | $\pm 5.1$ |
| total body fat [%] | | 29.5 | $\pm 7.4$ | | 25.9 | $\pm 8.4$ |
| Jump [cm] | | 146.3** | $\pm 42.5$ | | 172.5** | $\pm 46.2$ |
| handgrip [kg] | | 34.5 | $\pm 10.9$ | | 40.1 | $\pm 10.7$ |
| VO2max [ml O2/kg/min] | | 29.7* | $\pm 16.4$ | | 41.6* | $\pm 9.7$ |
| physical condition | 29 | -0.31* | $\pm 0.99$ | 29 | 0.29* | $\pm 0.97$ |
| Physical fitness | 29 | -0.26* | $\pm 0.89$ | 29 | 0.30* | $\pm 1$ |

\* $\leq .05$ ; \*\* $\leq .01$

### 2.3 Physical Fitness Analysis

To determine the participants’ physical fitness, they were asked to perform a fitness test, which was divided into a physical exercise part and body measurements, which resulted in variables representative for the participants’ physical appearance, such as Body Mass Index (BMI), waist-to-hip ratio (WHR), waist-to-height ratio (WHtR) and total body fat percentage. Body fat was assessed using a skinfold caliper on 7 anatomical points: triceps, subscapular, biceps, abdominal, suprailiac, thigh and calf. Skin was gripped between thumb and index finger. The caliper was applied about 1 cm from the fingers. To calculate the total body fat percentage after Siri (1961) and Brozek (1963) [36,37], body density was calculated first according to the age-specific formulas after Durnin and Womersley (1974) [38], using the sum of 4 skinfolds (triceps, biceps, subscapular, suprailiac).

The physical exercise part included a handgrip test to measure the muscular strength of the upper body. Participants were instructed to squeeze the Takei dynamometer (Takei Analogue Hand Grips Dynamo Meter, PS219A) as tightly as possible using one hand only and leaving their arms hanging next to their body with extended elbow joints. They performed this test twice with both hands at a time. A mean was calculated afterwards depicting the participants’ strengths in kilograms. To capture the muscular strength of the lower body, participants performed a standing long jump test. They were asked to line up behind a marker on the floor and to use their whole body dynamically in order to jump off using both legs simultaneously. They were specifically asked to land on both feet in the stand. Each participant had two attempts, of which the better one was noted down. The width of the jump was measured in centimeters, ranging from the marker to the back of the heel. The participants’ maximal oxygen capacity (VO2max) was determined using the Chester step test [39]. A pulse band was attached below the subjects’ chest and the pulse at rest was noted prior to the test. They were then instructed to step on the 30 cm-step moving one foot after the other - without jumping - at a step rate according to a metronome. The first level started with 60 beats per minute (bpm). The entire test consisted of 5 levels with each level being 2 minutes long. The metronome increased in speed with every level by 20 bpm, resulting in 140 bpm in level 5. The test was terminated as soon as the level was completed during which the subject had reached 80% of it’s age-related maximum heart rate (0.8 x (220 - age)) or if the subject felt too exhausted to continue. The Participants exhaustion was measured according to the Borg Rating of Perceived Exertion [40] in each level reached. The aerobic capacity was then manually estimated by plotting the exercise heart rates of each level completed on a prepared graphical datasheet. VO2max (ml O2/kg/min) could then be read off the **x**-axis, where the regression line intersected the line of maximum heart rate [39].

In order to include all variables acquired during the fitness test in the analysis but to reduce redundancy of the data at the same time, a principal component analysis (PCA) was performed. The 8 variables of the fitness test included in the PCA are listed in Table 4 of the supplementary section. As the factors were expected to be independent, a varimax rotation was used and item loadings below 0.30 were not considered. We furthermore chose two principal factors a priori, which then explained 75% of the data’s variance. The according scree plot can be can be found in Figure 2 and the factor loadings after rotation can be obtained from Table 4 of the supplementary section. As the correlation matrix was non-positive definite, which means that some of the eigenvalues are not positive values, the Kaiser-Meyer-Olkin measure and Bartlett test cannot be calculated. For further analysis, such as correlations, the two factors resulting from the PCA were used. We defined factor 1 as physical fitness and factor 2 as physical condition, describing the participants’ physical appearance. The values contained in factor 2 were recoded so that higher values can be interpreted as better physical condition.

**Figure 1.**
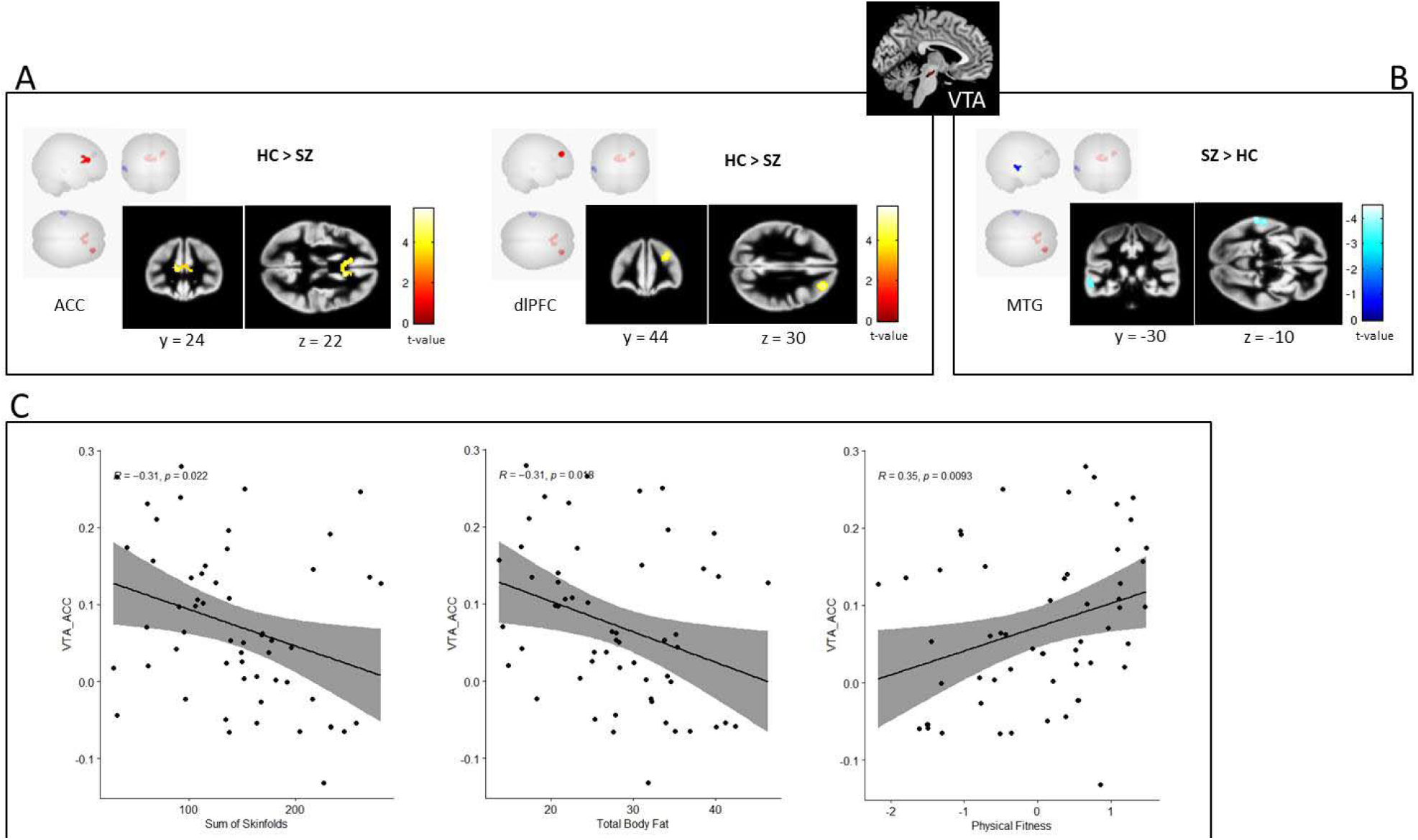
Seed-based connectivity results and correlations with physical fitness data. A shows two significant clusters resulting from the contrast healthy controls minus patients (HC > SZ) depicting increased functional connectivity between VTA (seed) and ACC (A, left) and increased functional connectivity between VTA and dlPFC (A, right) in the HC group compared to patients. B indicates increased functional connec tivity between VTA and MTG in the patient group compared to healthy controls (contrast SZ > HC). Color bars represent t-values (cluster threshold p < .05 FDR-corr.; voxel threshold p < .001 uncorr.). C shows significant correlations (p ≤ .05, uncorrected, for corrected values see Table 3) between the ACC cluster (A, left) and physical fitness parameters, including a negative correlation with the sum of skinfolds (C, left), a negative correlation with total body fat percentage (C, middle) and a positive correlation with overall physical fitness (C, right).

**Figure 2.**
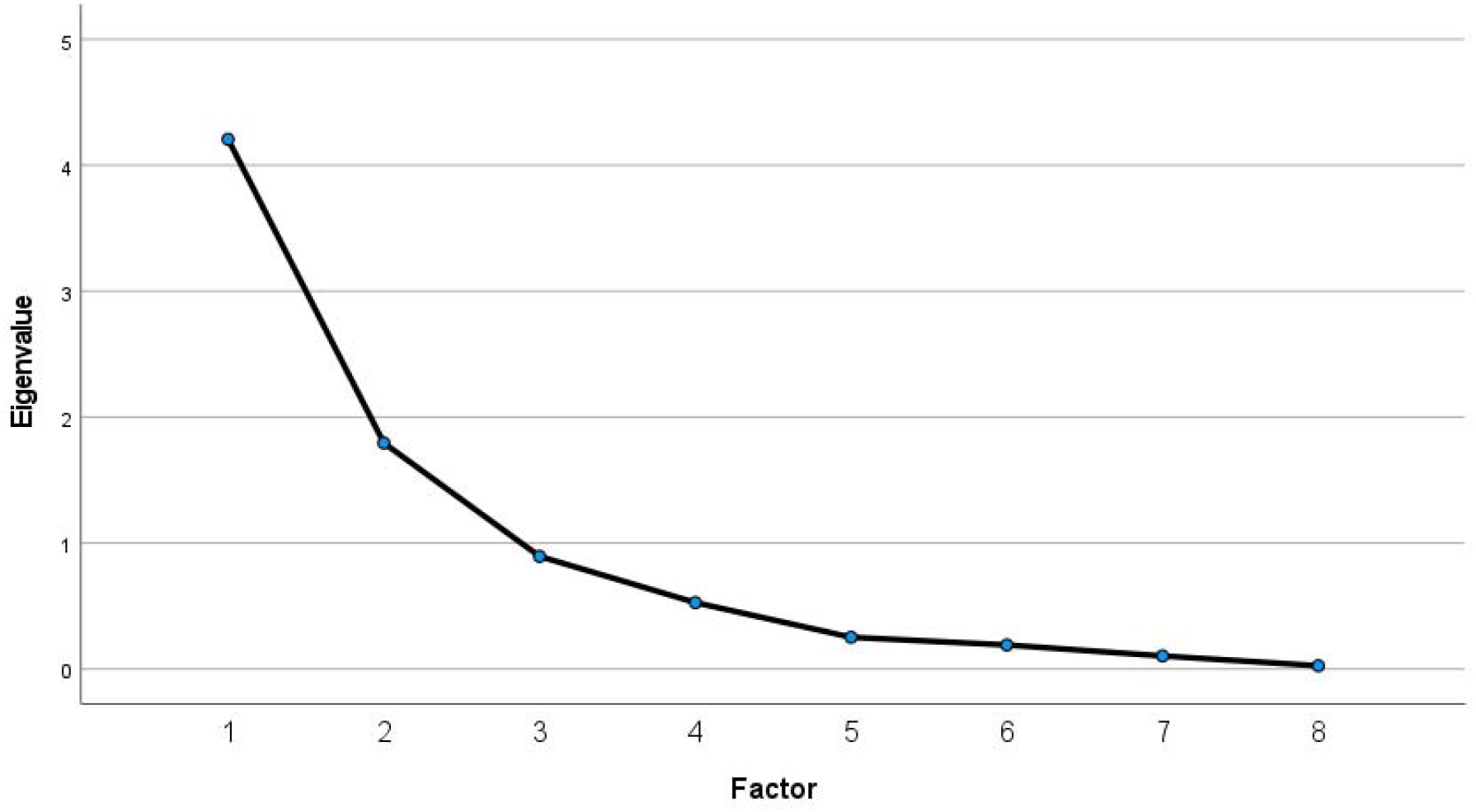
Screeplot of the principle component analysis. The x-axis displays the 8 factors, included in the PCA, resulting from the fitness test. The y-axis depicts the according eigenvalues of the factors and how much each factor contributes to explaining the data’s variance.

### 3.4 fMRI data acquisition and analysis

The functional MR images were obtained via a 3 Tesla MRI scanner (Siemens Magnetom Prisma) of the Brain Imaging Center of the Goethe University Frankfurt using a 64-channel head coil. The participants’ heads were stabilized with a small pillow belonging to the scanner equipment and equipped with ear plugs. They were instructed to rest quietly, focus on the crosshair shown on a display, visible through a mirror attached to the head coil, and not to fall asleep. The 262 volumes were acquired using a gradient-echo planar imaging (EPI) sequence with the following parameters: repetition time (TR) 1800 ms, echo time (TE) 30 ms, flip angle 90°, 52 transversal slices, slice thickness 2.8 mm, field of view (FoV) 224 mm (voxel size 2.8 × 2.8 × 2.8 mm). To compensate for distortions and to improve image quality, the EPI sequence was followed by a short field-map, with the same parameters consisting of only 3 images obtained in the opposite phase encoding direction - from posterior to anterior [41-43]. The according anatomical scans were obtained using a T1-weighted anatomical 3D magnetization-prepared rapid gradient echo sequence with the following parameters: TR = 2000 ms, TE = 2.12 ms, FoV = 256, 176 sagittal slices, voxel size = 1 × 1 × 1 mm, flip angle = 8°.

Prior to preprocessing, image distortions were corrected by the fieldmap images with fsl topup (https://fsl.fmrib.ox.ac.uk/fsl/fslwiki/topup). Preprocessing was then performed using the default preprocessing pipeline of the CONN toolbox (CONN-fMRI Functional Connectivity toolbox v20c, http://www.nitrc.org/projects/conn) including motion correction, slice time correction, bandpass filtering (0.008 Hz<f<0.9 Hz), spatial normalization, coregistration to anatomical scans, smoothing with an 8 mm Gaussian kernel and segmentation into grey matter, white matter and cerebrospinal fluid. We excluded three patients who exceeded the third quartile + 1.5 interquartile range in the quality assurance plots of maximum motion or maximum global BOLD signal change even after motion correction from further analysis.

In order to reveal seed-to-voxel connectivity differences between the patient- and healthy participant group, the second-level analysis was performed including age and gender as nuisance variables. To account for medication and movement effects, we further added chlorpromazine equivalent [44-50] and mean head motion as covariates into the model. The VTA seed region was predefined based on the Wolfgang Pauli Atlas [51]. The seed-based connectivity maps were computed using Fischer-transformed bivariate correlations between the seed region BOLD times series and the BOLD time series of each individual voxel of the brain [52].

To calculate correlations between the VTA-seed based connectivity results, psychopathology and physical fitness data, we extracted the mean beta-values from the voxels within significant clusters of each participant revealed by the second level analysis of CONN. We first examined whether the different variables were normally distributed using the Shapiro-Wilk-Test. We then calculated exploratory Pearson correlations, if the data was normally distributed, and Spearman correlations, if the data was not normally distributed.

## 3 Results

### 3.1 Sample characteristics

Sample characteristics can be taken from Table 1. Due to excessive head movement in the scanner we had to exclude three male patients from further analysis, resulting in a sample size of 29 patients (SZ) and 27 healthy controls (HC). Two sample t-tests revealed significant differences between both groups as to psychopathology. Patients obtained significantly higher scores in all questionnaires, including PAS (Wilcoxon *W* = 978.5; p = .012), SAS (Wilcoxon *W* = 963.5; p = .024), CDSS (Wilcoxon *W* = 975.5; p = .013) and WHODAS (Wilcoxon *W* = 1051; p ≤ .001). According to the fitness test, the healthy participants performed significantly better in the standing long jump test (t = 2.423; p = .018), as well as in the Chester step test (Wilcoxon *W* = 681.5; p = .017). The anthropometric measures further revealed lower BMIs for the healthy participants group, compared to the patients (Wilcoxon *W* = 965; p = .023). The groups did, however, not differ as to total body fat percentage (p = .068) and performed equally in the handgrip task (p = .064).

### 3.2 VTA-seed based connectivity results

The second level seed-to-voxel analysis of the resting-state data yielded three significant clusters, which are listed in Table 2 (cluster threshold p < .05 FDR-corr.; voxel threshold p < .001 uncorr.). Using the VTA as our seed, we found increased functional connectivity with the anterior cingulate cortex (ACC) and the right dorsolateral prefrontal cortex (dlPFC) in the healthy participant group compared to the patient group (contrast: HC > SZ). Patients showed increased functional connectivity between the VTA and the left middle temporal gyrus (MTG; HC < SZ). For visual presentation of the significant voxels in cortical areas only, we used a default grey matter mask provided by CONN to display the clusters (Figure 1).

**Table 2.** VTA seed-based functional connectivity results between patients and healthy controls.

| contrast | brain region | cluster size<br>[voxels] | peak MNI coordinates |  |  | peak t-value | p-value (FDR-corr.) |
| --- | --- | --- | --- | --- | --- | --- | --- |
|  |  |  | x | y | z |  |  |
| HC > SZ | anterior cingulate cortex | 293 | -5<br>7 | 27<br>30 | 18<br>17 | 4.95 | 0.0007 |
|  | dorsolateral prefrontal cortex, right | 174 | 33 | 45 | 30 | 4.42 | 0.007 |
| HC < SZ | middle temporal gyrus, left | 121 | -60 | -28 | -12 | -4.1 | 0.02 |

### 3.3 Correlation results

To associate psychopathology and physical fitness with VTA functional connectivity, we calculated correlations between the three significant clusters resulting from the fMRI analysis (Table 2; Figure 1) and the CDSS-, PAS-, SAS-, and WHODAS-score, as well as the variables acquired during the fitness test. We found significant negative correlations between the ACC cluster and total body fat (BF; p = .044, FDR-corr.), as well as the sum of all skinfolds, acquired with the caliper (p = .044, FDR-corr.). We further found a positive correlation between the ACC cluster and overall physical fitness (p = .036, FDR-corr.), a variable resulting from the PCA. We did not find any significant correlations with psychopathology. The correlation results are summarized in Table 3.

**Table 3.**
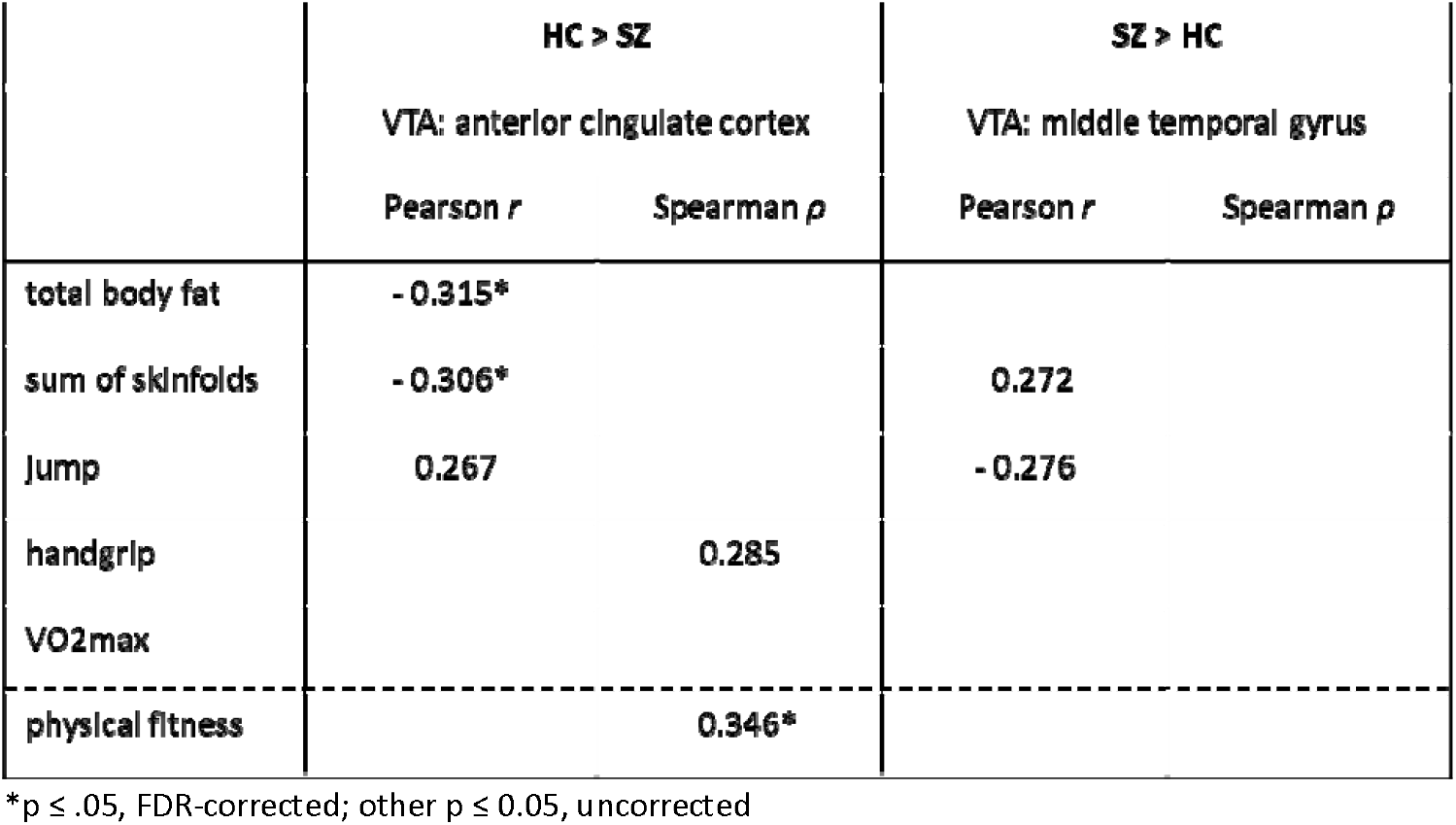
Significant correlation results between functional connectivity using VTA as a seed and physical fitness parameters.

**Table 4.** Rotated factor loadings after principle component analysis, of the fitness test parameters, using the varimax-method. We interpreted component 1 as physical fitness and component 2 as physical condition.

|  | Component 1 | Component 2 |
| --- | --- | --- |
| sum of skinfolds | -,651 | ,607 |
| BMI | -,325 | ,824 |
| BF | -,843 | ,355 |
| WHR |  | ,759 |
| WHtR | -,363 | ,906 |
| jump | ,887 |  |
| handgrip | ,836 | ,312 |
| VO2max | ,593 |  |

BF = total body fat percentage; WHR = waist-to-hip ratio; WHtR = waist to height ratio

## 4 Discussion

With this study, we wanted to verify whether dysfunctional connectivity patterns of the reward system are not only involved in the psychopathology of schizophrenia, but also associated with the decreased physical fitness, often observed in patients. To do so, we performed a seed-based functional connectivity analysis between the VTA and the rest of the brain and compared patients with controls. Finally, we correlated these outcomes with physical fitness data and psychopathology. Our results indicate decreased seed-based functional connectivity between the VTA and the dlPFC, as well as the VTA and ACC in our group of patients with schizophrenia compared to healthy participants. We further found increased functional connectivity between the VTA and the MTG in the patient group. Moreover, the decreased functional connectivity between the VTA and the ACC could be associated with elevated body fat. On the other hand, higher functional connectivity between the VTA and the ACC correlated positively with overall physical fitness. These findings support our original hypothesis that striatal dysconnectivity in schizophrenia is linked to decreased physical fitness.

Our findings are in line with previous studies, demonstrating decreased physical fitness in patients with schizophrenia [53]. In our sample, patients have significantly higher BMIs than the healthy controls and performed significantly worse in the Chester step test, measuring cardiorespiratory fitness, as well as the standing long jump test, compared to healthy controls. The PCA further yielded two variables, one reflecting the overall physical condition (or appearance), the other representing the overall physical fitness. Patients obtained significantly lower values as to both parameters. This impaired physical fitness of patients has many causes, especially associated with the mental illness itself and is closely linked to their increased risk of weight gain. Besides the identified genetic predisposition, there are environmental and lifestyle factors, which render patients more susceptible to gaining weight. These are social stigmatization and discrimination due to mental illness, social isolation and poor symptom management, increasing sedentary behavior, the lack of motivation to exercise and food intake [54-56]. Furthermore, there have been some interesting speculations about how the ACC is involved in non-exercise physical activity [57]. While most studies focus on the effects of exercise on mental well-being, there have been only a few studies investigating the impact of non-exercise everyday life activity on mental health. However, according to Reichert and colleagues (2020), increased physical activity, comprising all physical activities performed in everyday-life, are associated with higher grey matter volumes of the ACC, more specifically the subgenual part. This might explain why we have found increased physical activity to correlate with stronger functional connectivity between the VTA and the ACC.

Up to now, only a few studies have investigated functional connectivity of the VTA in schizophrenia, which makes comparisons and interpretations of results somewhat difficult. Yet, our resting state outcomes are consistent with the findings of Hadley and colleagues [58], who found a functional connection between the VTA and the ACC, as well as the inferior frontal gyrus in healthy participants. Structural and functional impairments of the anterior cingulate cortex could be associated with the psychopathology of schizophrenia. There is evidence that especially negative symptomatology correlates with the functional dysregulation of the ACC, which is implicated in emotional and cognitive processes as well as goal-directed behavior [59-61]. However, the decreased functional connectivity between the VTA and the ACC in our sample of schizophrenic patients was not directly related to negative symptomatology. Interestingly, there is some evidence that the urban social environment and upbringing affects social-emotional processing of limbic circuits involving the ACC as a key node [62]. As we did not include information about the social environment or upbringing of our participants as a variable in the analysis of this study, we can only speculate if the decreased functional connectivity between the VTA and the ACC in our patient group is linked to social influential factors. This should therefore be taken into account in future analyses.

To our knowledge, we are the first to identify decreased functional connectivity between the VTA and the dlPFC in schizophrenia patients. Several studies have characterized the role of the dlPFC in terms of cognitive impairments, especially working memory deficits, in patients [63-65]. Thus a decreased functional connectivity between the VTA and the dlPFC in schizophrenia patients compared to healthy controls seems plausible as it might reflect the patients’ cognitive disabilities. However, further research will be necessary to support this hypothesis, since we did not explicitly assess cognition in this study.

We have further found stronger functional connections between the VTA and the left MTG in the patient group. The left MTG has been moved into focus as it could be related to auditory and verbal hallucinations in the past [66] and shows reductions in grey matter volume that have been associated with the psychopathology [67]. To address the question whether the increased functional connectivity between the VTA and the MTG is a result of an overcompensation due to its reduction in size or linked to psychosis is speculative. There are, however, a few studies, which have found increased functional connections between the striatum and the middle temporal gyrus [68]. Future resting-state studies using a VTA-seed should give further insights.

Since the dopaminergic reward system regulates motivational behavior, including voluntary movement and eating behavior [69,70], we assumed that the reward system represents the link between decreased physical fitness and psychopathology in schizophrenia. Further arguments supporting this assumption are based on the fact that pathological changes in functional connectivity of the reward system could be associated with obesity [71]. Our results support this hypothesis by significant correlations between physical fitness parameters and the functional connectivity of the VTA and ACC. Follow up analysis should additionally include comparisons between patients and obese, but otherwise healthy, individuals, in order to discover possible similarities within connectivity patterns.

This study faces a few limitations. First, although there is a great body of evidence that striatal dysconnectivity correlates with symptom severity in schizophrenia, we did not find significant correlations between VTA-seed based functional connections and psychopathology, neither within the whole study sample, nor within the patient group only. This could, on the one hand, be due to our relatively small sample size, and, on the other hand, because of the fact, that our psychopathological assessment is mostly based on self-evaluation, requiring concentration and disease insight. Second, our fitness test data could be biased by the fact that antipsychotics can impact pulse regulation via anticholinergic effects. These effects on the Chester Step test have not been systematically assessed. Moreover, antipsychotic medication plays an important role in weight gain of schizophrenia [72]. But as medication effects can only be reliably assessed in longitudinal designs, we used it as a covariate to adjust for. Apart from that, many studies have demonstrated decreased physical fitness and weight gain also in drug-naïve patients []. Third, in contrast to task-based paradigms, resting state fMRI measurements cannot control the participants’ thinking as it is the case in clearly operationalized tasks. Finally, our study is a cross-sectional study, which does not give insights into long-term effects of physical exercise interventions. In future longitudinal studies it would be interesting to see how the VTA-seed based functional connections might be altered by lifestyle therapies.

In summary, our findings provide deeper insights into less frequently addressed, but no less burdensome comorbidities of schizophrenia. We could demonstrate that decreased physical fitness in schizophrenia is linked to altered functional connectivity of the VTA. Excessive weight gain and decreased physical fitness pose a major problem, as they increase the disease burden, lower self-esteem, as well as the patients’ life expectancies. Thus, besides the treatment of the negative symptoms, the focus in schizophrenia research needs to be shifted more towards the consequences on physical health. As a brain circuit which regulates behavioral responses, the reward system represents a suitable target for behavioral interventions in the future, including lifestyle therapies and physical exercise.

## Data Availability

All data produced in the present study are available upon reasonable request to the authors

## Supplements

### 1 Principle component analysis results

## References

1. Freedman R. Schizophrenia. N Engl J Med. 18, 1738–49; 10.1056/NEJMra035458 (2003).

2. American Psychiatric Association. Diagnostic and statistical manual of mental disorders (5th ed.). https://doi-org.ezproxy.frederick.edu/10.1176/appi.books.9780890425596 (2013).

3. Charlson, F. J. et al. Global epidemiology and burden of schizophrenia. Findings from the global burden of disease study 2016. Schizophrenia Bulletin 44, 1195–1203; 10.1093/schbul/sby058 (2018).

4. Bak, M., Fransen, A., Janssen, J., van Os, J. & Drukker, M. Almost all antipsychotics result in weight gain. A meta-analysis. PLoS ONE 9, 10–12; 10.1371/journal.pone.0094112 (2014).

5. Strassnig, M., Brar, J. & Ganguli, R. Low cardiorespiratory fitness and physical functional capacity in obese patients with schizophrenia. Schizophrenia Research 126, 103–109; 10.1016/j.schres.2010.10.025 (2010).

6. Laaksonen, D. E. et al. Low levels of leisure-time physical activity and cardiorespiratory fitness predict development of the metabolic syndrome. Diabetes Care 25, 1612–1618; 10.2337/diacare.25.9.1612 (2002).

7. LaMonte, M. J. et al. Cardiorespiratory fitness is inversely associated with the incidence of metabolic syndrome. A prospective study of men and women. Circulation 112, 505–512; 10.1161/CIRCULATIONAHA.104.503805 (2005).

8. Thakore, J. Metabolic syndrome and schizophrenia. British Journal of Psychiatry, 186(6), 455–456; 10.1192/bjp.186.6.455 (2005).

9. Ford, E. S. & Li, C. Physical activity or fitness and the metabolic syndrome. Expert Review of Cardiovascular Therapy 4, 897–915; 10.1586/14779072.4.6.897 (2006).

10. Grajales, D., Ferreira, V. & Valverde, Á. Second-Generation Antipsychotics and Dysregulation of Glucose Metabolism. Beyond Weight Gain. Cells 8, 1336; 10.3390/cells8111336 (2019).

11. Ratliff, J. C., Palmese, L. B., Reutenauer, E. L., Srihari, V. H. & Tek, C. Obese Schizophrenia Spectrum Patients Have Significantly Higher 10-Year General Cardiovascular Risk and Vascular Ages than Obese Individuals without Severe Mental Illness. Psychosomatics 54, 67–73; 10.1016/j.psym.2012.03.001 (2013).

12. Laursen, T. M., Munk-Olsen, T. & Vestergaard, M. Life expectancy and cardiovascular mortality in persons with schizophrenia. Current Opinion in Psychiatry 25, 83–88; 10.1097/YCO.0b013e32835035ca (2012).

13. Manu, P. et al. Weight gain and obesity in schizophrenia. Epidemiology, pathobiology, and management. Acta Psychiatrica Scandinavica 132, 97–108; 10.1111/acps.12445 (2015).

14. Elman, I., Borsook, D. & Lukas, S. E. Food intake and reward mechanisms in patients with schizophrenia. Implications for metabolic disturbances and treatment with second-generation antipsychotic agents. Neuropsychopharmacology 31, 2091–2120; 10.1038/sj.npp.1301051 (2006).

15. Howes, O. D. & Kapur, S. The dopamine hypothesis of schizophrenia. Version III - The final common pathway. Schizophrenia Bulletin 35, 549–562; 10.1093/schbul/sbp006 (2009).

16. Simpson, E. H., Kellendonk, C., & Kandel, E. A possible role for the striatum in the pathogenesis of the cognitive symptoms of schizophrenia. Neuron, 65(5), 585–596; 10.1016/j.neuron.2010.02.014 (2010).

17. McCutcheon, R. A., Reis Marques, T. & Howes, O. D. Schizophrenia - An Overview. JAMA Psychiatry 77, 201–210; 10.1001/jamapsychiatry.2019.3360 (2020).

18. Conn, K. A., Burne, T. H. & Kesby, J. P. Subcortical Dopamine and Cognition in Schizophrenia. Looking Beyond Psychosis in Preclinical Models. Frontiers in Neuroscience 14, 1–18; 10.3389/fnins.2020.00542 (2020).

19. Cohen, J. Y., Haesler, S., Vong, L., Lowell, B. B. & Uchida, N. Neuron-type-specific signals for reward and punishment in the ventral tegmental area. Nature 482, 85–88; 10.1038/nature10754 (2012).

20. Lammel, S. et al. Input-specific control of reward and aversion in the ventral tegmental area. Nature 491, 212–217; 10.1038/nature11527 (2012).

21. Meye, F. J. & Adan, R. A. Feelings about food. The ventral tegmental area in food reward and emotional eating. Trends in Pharmacological Sciences 35, 31–40; 10.1016/j.tips.2013.11.003 (2014).

22. Radua, J. et al. Ventral striatal activation during reward processing in psychosis a neurofunctional meta-analysis. JAMA Psychiatry 72, 1243–1251; 10.1001/jamapsychiatry.2015.2196 (2015).

23. Grimm, O. et al. Striatal response to reward anticipation evidence for a systems-level intermediate phenotype for schizophrenia. JAMA Psychiatry 71, 531–539; 10.1001/jamapsychiatry.2014.9 (2014).

24. Fornito, A. et al. Functional dysconnectivity of corticostriatal circuitry as a risk phenotype for psychosis. JAMA Psychiatry 70, 1143–1151; 10.1001/jamapsychiatry.2013.1976 (2013).

25. Martino, M. et al. Abnormal resting-state connectivity in a substantia nigra-related striato-thalamo-cortical network in a large sample of first-episode drug-naïve patients with schizophrenia. Schizophrenia Bulletin 44, 419–431; 10.1093/schbul/sbx067 (2018).

26. Cota, D., Barrera, J. G. & Seeley, R. J. Leptin in Energy Balance and Reward. Two Faces of the Same Coin? Neuron 51, 678–680; 10.1016/j.neuron.2006.09.009 (2006).

27. Abizaid, A. Ghrelin and dopamine. New insights on the peripheral regulation of appetite. Journal of Neuroendocrinology 21, 787–793; 10.1111/j.1365-2826.2009.01896.x (2009).

28. Woodworth, H. L. et al. Neurotensin Receptor-1 Identifies a Subset of Ventral Tegmental Dopamine Neurons that Coordinates Energy Balance. Cell Reports 20, 1881–1892; 10.1016/j.celrep.2017.08.001 (2017).

29. Beeler, J. A., Frazier, C. R. & Zhuang, X. Putting desire on a budget. Dopamine and energy expenditure, reconciling reward and resources. Frontiers in Integrative Neuroscience 6, 1–22; 10.3389/fnint.2012.00049 (2012).

30. Chapman, L. J., Chapman, J. P. & Raulin, M. P. Chapman, L.J., Chapman, J.P., & Raulin, M.L. (1976) Scales for physical and social anhedonia.pdf (1976).

31. Üstün, T. B. Measuring health and disability. Manual for WHO Disability Assessment Schedule WHODAS 2.0 (World Health Organization, Geneva, 2010).

32. Addington, D., Addington, J. & Maticka-Tyndale, E. Assessing depression in schizophrenia. The Calgary depression scale. British Journal of Psychiatry 163, 39–44; 10.1192/s0007125000292581 (1993).

33. Kay, S. R., Fiszbein, A. & Opler, L. A. The positive and negative syndrome scale (PANSS) for schizophrenia. Schizophrenia Bulletin 13, 261–276; 10.1093/schbul/13.2.261 (1987).

34. Mann, H. B. & Whitney, D. R. On a Test of Whether one of Two Random Variables is Stochastically Larger than the Other. The Annals of Mathematical Statistics 18, 50–60; 10.1214/aoms/1177730491 (1947).

35. Shapiro, S. & Wilk, M. B. An Analysis of Variance Test for Normality (Complete Samples) Published by. Biometrika Trust Stable URL : http://www.jstor.org/stable/2333709. Biometrika 52, 591–611 (1965).

36. Siri, W. E. & Lukaski, H. C. Body composition from fluid spaces and density. Analysis of methods. Prospective Overview. Nutrition 9 (1993).

37. Brožek, J., Grande, F., Anderson, J. & Keys, A. Densitometric Analysis of Body Composition. Revision of Some Quantitative Assumptions. Annals of the New York Academy of Sciences 110, 113–140; 10.1111/j.1749-6632.1963.tb17079.x (2006).

38. Durnin, B. Y. J. V. G. a. & Womersley, J. and Its Estimation From Skinfold Thickness. Measurements on. British Journal of Nutrition 32, 77–97 (1973).

39. Sykes, K. & Roberts, A. The Chester step test-a simple yet effective tool for the prediction of aerobic capacity. Physiotherapy 90, 183–188; 10.1016/j.physio.2004.03.008 (2004).

40. Borg G. Perceived exertion as an indicator of somatic stress. Scand J Rehabil Med 2(2); 92–8; PMID: 5523831 (1970).

41. Andersson, J. L., Skare, S. & Ashburner, J. How to correct susceptibility distortions in spin-echo echo-planar images. Application to diffusion tensor imaging. NeuroImage 20, 870–888; 10.1016/S1053-8119(03)00336-7 (2003).

42. Smith, S. M. et al. Advances in functional and structural MR image analysis and implementation as FSL. NeuroImage 23, 208–219; 10.1016/j.neuroimage.2004.07.051 (2004).

43. Shrestha, M., Hok, P., Nöth, U., Lienerth, B. & Deichmann, R. Optimization of diffusion-weighted single-refocused spin-echo EPI by reducing eddy-current artifacts and shortening the echo time. Magnetic Resonance Materials in Physics, Biology and Medicine 31, 585–597; 10.1007/s10334-018-0684-x (2018).

44. Woods, S. W. Chlorpromazine equivalent doses for the newer atypical antipsychotics. Journal of Clinical Psychiatry 64, 663–667; 10.4088/JCP.v64n0607 (2003).

45. Schatzberg A.F., Cole J.O. & DeBattista C. Manual of Clinical Psychopharmacology. American Psychiatric Publishing 7th Edition (2010).

46. Taylor D., Paton C., & Kapur S. The Maudsley Prescribing Guidelines in Psychiatry, Wiley-Blackwell; 11th Edition (2011).

47. Patel, M. X., Arista, I. A., Taylor, M. & Barnes, T. R. How to compare doses of different antipsychotics. A systematic review of methods. Schizophrenia Research 149, 141–148; 10.1016/j.schres.2013.06.030 (2013).

48. Leucht, S. et al. Dose equivalents for second-generation antipsychotics. The minimum effective dose method. Schizophrenia Bulletin 40, 314–326; 10.1093/schbul/sbu001 (2014).

49. Leucht, S. et al. Dose Equivalents for Second-Generation Antipsychotic Drugs. The Classical Mean Dose Method. Schizophrenia Bulletin 41, 1397–1402; 10.1093/schbul/sbv037 (2015).

50. Rothe P.H., Heres S. & Leucht S. Dose equivalents for second generation long-acting injectable antipsychotics: The minimum effective dose method. Schizophrenia Res. 193, 23–28; 10.1016/j.schres.2017.07.033 (2018).

51. Pauli, W. M., Nili, A. N. & Michael Tyszka, J. Data Descriptor. A high-resolution probabilistic in vivo atlas of human subcortical brain nuclei. Scientific Data 5, 1–14; 10.1038/sdata.2018.63 (2018).

52. Nieto-Castanon, A. Handbook of functional connectivity Magnetic Resonance Imaging methods in CONN (2020).

53. Ozbulut, O. et al. Evaluation of physical fitness parameters in patients with schizophrenia. Psychiatry Research 210, 806–811; 10.1016/j.psychres.2013.09.015 (2013).

54. Thakore, J. H., Mann, J. N., Vlahos, I., Martin, A. & Reznek, R. Increased visceral fat distribution in drugnaive and drug-free patients with schizophrenia. International Journal of Obesity 26, 137–141; 10.1038/sj.ijo.0801840 (2002).

55. Palmese, L., Reutenauer, E., Liskov, E., Grilo, C. & Tek, C. The effect of dietary and physical activity pattern on metabolic profile in individuals with schizophrenia. A cross-sectional study. Comprehensive psychiatry 53, 1028–1033; 10.1016/j.comppsych.2012.02.003 (2012).

56. Annamalai, A., Kosir, U. & Tek, C. Prevalence of obesity and diabetes in patients with schizophrenia. World Journal of Diabetes 8, 390; 10.4239/wjd.v8.i8.390 (2017).

57. Reichert, M. et al. A neural mechanism for affective well-being. Subgenual cingulate cortex mediates real-life effects of nonexercise activity on energy. Science Advances 6; 10.1126/SCIADV.AAZ8934 (2020).

58. Hadley, J. A. et al. Ventral tegmental area/midbrain functional connectivity and response to antipsychotic medication in schizophrenia. Neuropsychopharmacology 39, 1020–1030; 10.1038/npp.2013.305 (2014).

59. Dolan R.J. et al. Dopaminergic modulation of impaired cognitive activation in the anterior cingulate cortex in schizophrenia. Nature 378(6553), 180–2; 10.1038/378180a0 (1995).

60. Fornito, A., Yücel, M., Dean, B., Wood, S. J. & Pantelis, C. Anatomical abnormalities of the anterior cingulate cortex in schizophrenia. Bridging the gap between neuroimaging and neuropathology. Schizophrenia Bulletin 35, 973–993; 10.1093/schbul/sbn025 (2009).

61. Nelson B.D., Bjorkquist O.A., Olsen E.K. & Herbener E.S. Schizophrenia symptom and functional correlates of anterior cingulate cortex activation to emotion stimuli: An fMRI investigation. Psychiatry Res. 234(3), 285–91; 10.1016/j.pscychresns.2015.11.001 (2015).

62. Tost, H., & Meyer-Lindenberg, A. Puzzling over schizophrenia: Schizophrenia, social environment and the brain. Nature Medicine, 18, 211–213,10.1038/nm.2671 (2012).

63. Bähner, F. & Meyer-Lindenberg, A. Hippocampal–prefrontal connectivity as a translational phenotype for schizophrenia. European Neuropsychopharmacology 27, 93–106; 10.1016/j.euroneuro.2016.12.007 (2017).

64. Huang, M. L. et al. Relationships between dorsolateral prefrontal cortex metabolic change and cognitive impairment in first-episode neuroleptic-naive schizophrenia patients. Medicine (United States) 96; 10.1097/MD.0000000000007228 (2017).

65. Smucny, J., Dienel, S. J., Lewis, D. A. & Carter, C. S. Mechanisms underlying dorsolateral prefrontal cortex contributions to cognitive dysfunction in schizophrenia. Neuropsychopharmacology 47, 292–308; 10.1038/s41386-021-01089-0 (2022).

66. Zhang, L. et al. Decreased middle temporal gyrus connectivity in the language network in schizophrenia patients with auditory verbal hallucinations. Neuroscience Letters 653, 177–182; 10.1016/j.neulet.2017.05.042 (2017).

67. Guo, W. et al. Decreased gray matter volume in the left middle temporal gyrus as a candidate biomarker for schizophrenia. A study of drug naive, first-episode schizophrenia patients and unaffected siblings. Schizophrenia Research 159, 43–50; 10.1016/j.schres.2014.07.051 (2014).

68. Zhou B., Tan C., Tang J. & Chen X. Brain functional connectivity of functional magnetic resonance imaging of patients with early-onset schizophrenia. Zhong Nan Da Xue Xue Bao Yi Xue Ban 35(1), 17–24; 10.3969/j.issn.1672-7347.2010.01.003 (2010).

69. Beeler, J. A., Frazier, C. R. & Zhuang, X. Putting desire on a budget. Dopamine and energy expenditure, reconciling reward and resources. Frontiers in Integrative Neuroscience 6, 1–22; 10.3389/fnint.2012.00049 (2012).

70. Mourra D., Gnazzo F., Cobos S. & Beeler J.A. Striatal Dopamine D2 Receptors Regulate Cost Sensitivity and Behavioral Thrift. Neuroscience 425, 134–145; 10.1016/j.neuroscience.2019.11.002 (2020).

71. Park, B.-Y. et al. Whole-brain functional connectivity correlates of obesity phenotypes. Human Brain Mapping 41; 10.1002/hbm.25167 (2020).

72. Homan, P. et al. Striatal volume and functional connectivity correlate with weight gain in early-phase psychosis. Neuropsychopharmacology 44, 1948–1954; 10.1038/s41386-019-0464-y (2019).

73. Thakore, J. H., Mann, J. N., Vlahos, I., Martin, A. & Reznek, R. Increased visceral fat distribution in drug-naive and drug-free patients with schizophrenia. International journal of obesity and related metabolic disorders: journal of the International Association for the Study of Obesity 26, 137–141; 10.1038/sj.ijo.0801840 (2002).

